# A Deep Learning Enhanced Compartmental Model and its Application in Modeling Omicron Dynamics and Development in China

**DOI:** 10.1101/2022.06.05.22276023

**Authors:** Qi Deng

**Author notes:** corresponding author: address: 167 West Checheng Road, Shiyan, Hubei 442002, China.

## Abstract

The mainstream compartmental models require stochastic parameterization to estimate the transmission parameters between compartments, which depends upon detailed statistics on epidemiological characteristics that are economically and resource-wide expensive to collect. As an alternative, deep learning techniques are effective in estimating these stochastic parameters with greatly reduced dependency on data particularity. We apply deep learning to estimate transmission parameters of a customized compartmental model, then feed the estimated transmission parameters to the compartmental model to predict the development of the Omicron epidemics in China for 28 days. The average levels of predication accuracy of the model are 98% and 92% for number of infections and deaths, respectively.

## 1. Introduction

The Omicron variant of COVID-19 started invading China as early as November 2021 and became a full-fledge epidemic in late February 2022. Assuming that all confirmed infections since November 31, 2022, are Omicron cases, as of June 3, 2022, there have been 764,369 confirmed Omicron cases, and 590 patients have died with Omicron infection in mainland China. The numbers of confirmed and deceased cases are 1,214,192 and 9,382, respectively, in Hong Kong. For Taiwan, the numbers are 2,274,666 and 2,663.

Most of the research on Omicron focuses on the effectiveness of immunization, vaccination and treatment, with relatively few epidemiological studies on the variant, especially in a Chinese context. A computational simulation-model study utilizing a customized Monte Carlo model to estimate the effect of facemask use before and after different COVID-19 vaccination coverage levels was conducted [1]. A set of posterior statistical models to estimate cumulative infections and cumulative proportion of the populations in worldwide locations is produced [2]. While epidemiological studies on Omicron are lacking in general, there have been attempts to model the original variant dynamics in China. A classic SEIR model is used to infer the basic reproduction ratio and to simulate the Wuhan epidemic [3]. More sophisticated models have been developed to correlate the risk levels of foreign countries with their travel exposure to China [4, 5], including a stochastic dual-SEIR approach on both the Wuhan population and international travelers to estimate how transmission has varied over time from Wuhan to international destinations [5]. Simulations on international spread after the start of travel ban from Wuhan on Jan 23, 2020 have also been conducted [6], which apply the Global Epidemic and Mobility Model (GLEAM) to a multitude of Chinese and global cities and a SEIR variety (SLIR) to project the impact of human-to-human transmissions.

Since March 2020, with the outbreak of the original variant declining in China, researchers have dedicated more efforts to analyzing the effectiveness of containment measures. Mobility and travel history data from Wuhan are used to ascertain the impact of the drastic control measures implemented in China [7]. The spread and control of COVID-19 among Chinese cities with data on human movements and public health interventions has been investigated [8]. A transmission model to study the impact of social distancing and school closure is built, utilizing the contact data for Wuhan and Shanghai and tracing information from Hunan [9].

In late February 2022, China, especially Shanghai, was hit hard by the Omicron variant. With a much higher initial reproduction rate than that of the original and Delta variants at between 1.72 and 8.2 [10, 11, 12], the Omicron variant presents a potentially game-changing challenge to the country’s zero-COVID strategy that was effective against the original and Delta variants. An age-structured stochastic compartmental model (SLIRS) calibrated on the initial growth phase for the 2022 Omicron outbreak in Shanghai is developed [13]. The key contribution of the model is the inclusion of age-specific vaccine coverage data, vaccine efficacy against different clinical endpoints, waning of immunity, different antiviral therapies, and nonpharmaceutical interventions.

The aim of this paper is to establish a class of innovative compartmental models, of which the transmission parameters are estimated by a family of multivariate, multistep deep learning methodologies. The models are then used to predict and simulate the dynamics of Omicron in China (both including and excluding Hong Kong and Taiwan), as well as Hong Kong and Taiwan, for a 28-day period between June 3 and July 1, 2022. We then compare the reported numbers of infections and deaths to measure the prediction accuracy of our models. The average levels of prediction accuracy provided by the models are 98% and 92% for the numbers of infections and deaths, respectively.

## 2. Materials and Methods

### 2.1 Compartmental Models

The mainstream compartmental models require detailed statistics on the characteristics of an infectious disease to estimate the stochastic transmission parameters between compartments. Essentially, these models correlate tempo-spatial factors (e.g., geographic distances and contact durations) among heterogeneous subpopulations with gradient probability decays. Theoretically, carefully designed transmission parameterization processes utilize Bayesian inference methods, such as Marcov Chain Monte Carlo (MCMC) or Gillespie algorithm [14] simulations, to form probability density functions (PDFs) on cross-sections to estimate transmission parameters for each timestep of a compartmental time series, which need further calibration with historical transmission data to achieve a reasonable level of accuracy. However, detailed statistics on transmission characteristics are economically and resource-wise expensive to collect. As an alternative, some researchers [e.g., 13] simply assume the values of these transmission parameters to achieve cost-effectiveness.

We are particularly interested in compartmental models that cover multiple interconnected and heterogeneous subpopulations [9, 15, 16]. We first develop a multistep, multivariate deep learning methodology to estimate the transmission parameters and then feed them to a class of customized compartmental models to predict and simulate the development of the Omicron epidemic in China (including and excluding Hong Kong and Taiwan), Hong Kong and Taiwan.

We establish a SIR-derived discrete time series on a daily interval as the theoretical foundation for a deep learning-enhanced compartment model – SIRD (Susceptible-Infectious-Removed-Deceased). A precursor to this study is developed to predict and simulate the dynamics and development of the original COVID-19 variant in the US [17].

The SIRD construct groups a population into four compartments:

1. Susceptible (S): The susceptible population that progresses into the infectious compartment.
2. Infectious (I): The infectious individuals who are symptomatic come from the Susceptible compartment and progress into the Removed compartment.
3. Removed (R): The recovered individuals come from the infectious compartment and acquire lasting immunity (there has yet to be any contradiction against this assumption for Omicron).
4. Deceased (D): The deceased cases come from the infectious compartment.

The SIRD model has a discrete daily (Δ*t = 1*) multivariate time series construct given by the following matrix form:

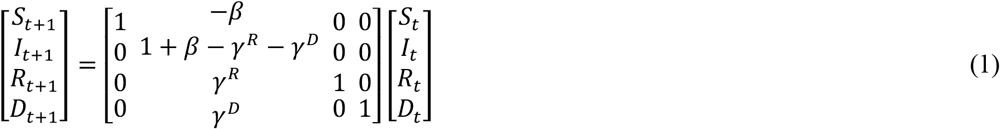

Or:

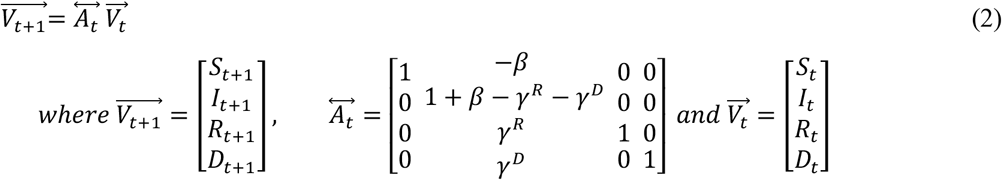

The Greek letters *β, γ*^*R*^, *γ*^*D*^ *i*n Equations 1 and 2 are the “susceptible-to-infectious,” “infectious-to-removed,” and “infectious-to-deceased” transmission parameters.

Since we need to estimate the transmission parameters, we rewrite and rearrange Equations 1 and 2 to the following matrix representation:

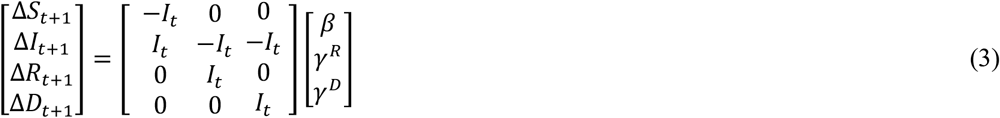

or:

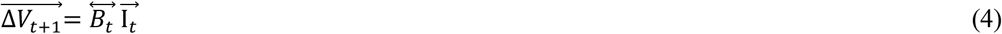

*Where*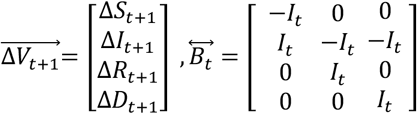 *and* 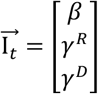

### 2.2 Parameterization with Deep Learning and SIRD Simulation

The transmission parameters (*β, γ*^*R*^, *γ*^*D*^) in Equations 1 to 4 are both non-stochastic values in the temporal dimension (*t*) and stochastic variables along three “spatial dimensions,” namely, population distribution (*S*), population mobility (*L*), and population heterogeneity (*C*). A parameterization to estimate the transmission parameters at each time step (cross-section in the multivariate SIRD construct) is therefore required and has the following expression:

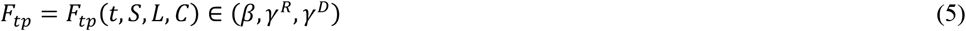

Equation 5 shows that each transmission parameter ( *F*_*tp*_) can be modeled in a 4-dimensional tempo-spatial framework. The parameterization process is thus to estimate the in-sample values of cross-sectional *F*_*tp*_ at each time step *t* in the SIRD time series construct and predict its out-of-sample values.

We aim to build a multistep, multivariate deep learning method to estimate the transmission parameters, utilizing both the standard deep neural network (DNN) and the advanced recurrent neural network – long short-term memory neural network (RNN-LSTM, or simply LSTM) methodologies. We propose the following steps to achieve this goal:

1. Constructing the in-sample SIRD time series using observed Omicron data.
2. Calculating in-sample daily transmission parameters from the in-sample SIRD time series constructed in Step 1.
3. Decomposing Equation (5) as: *F*_*tp*_ *= F*_*tp*_(*t, S, L, C*) *= F*_*tp*_(*t*)Ψ(*S, L, C*). That is, along the temporal dimension, at the given time step *t*, the nonstochastic value of the transmission parameter is *F*_*tp*_(*t*); along the spatial dimensions (S, L, C), the cross-sectional probability distribution of the transmission parameter is Ψ(*S, L, C*).
4. Deep learning algorithms (DNN and LSTM) are applied to fit the in-sample decomposed transmission parameters in step 3. Deep learning is performed on both *F*_*tp*_(*t*) *a*nd Ψ(*S, L, C*) *t*o calibrate the in-sample values of *F*_*tp*_(*t, S, L, C*) *a*long both temporal and spatial dimensions, respectively.
5. With the in-sample transmission parameters obtained in step 4, the DNN and RNN-LSTM algorithms are applied again, in both progressive and recursive manners, to predict the out-of-sample transmission parameters for multiple scenarios.
6. Simulating out-of-sample Omicron dynamics recursively through the SIRD time series, using the out-of-sample transmission parameters predicted in step 5.

The methodological innovation of our research is mainly reflected in deep learning of the cross-sectional probability distribution Ψ(*S, L, C*). The DNN decomposition of the parameter *f*_*tp*_ is expressed as:

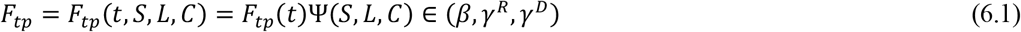

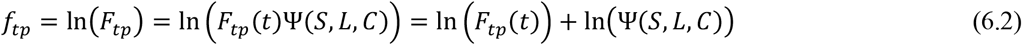

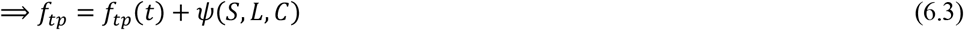

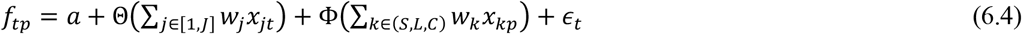

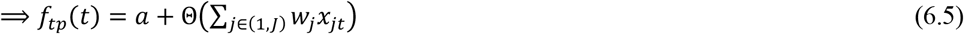

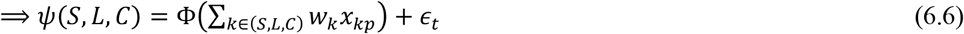

For in-sample data, *f*_*tp*_ *i*s the actual (observed) transmission parameter value (logarithmic) of the SIRD time series, *f*_*tp*_(*t*) *i*s the non-stochastic parameter value (logarithmic) in the temporal dimension, and *ψ*(*S, L, C*) *i*s the cross-sectional probability distribution in the spatial dimensions (logarithmic). The importance of decomposing *f*_*tp*_ *i*nto *f*_*tp*_(*t*) *a*nd *ψ*(*S, L, C*) *i*s to independently calibrate the temporal non-stochastic value and the spatial probability distribution of *f*_*tp*_ *t*o further estimate its out-of-sample values.

The LSTM decomposition of *f*_*tp*_ *i*s expressed as:

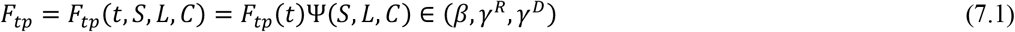

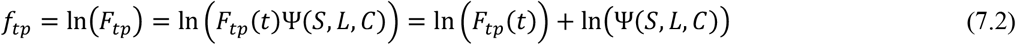

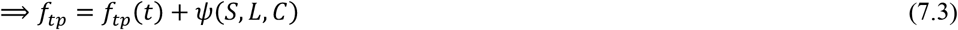

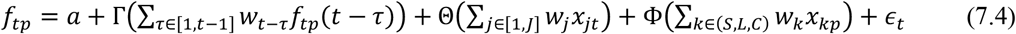

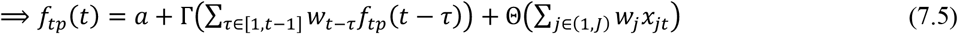

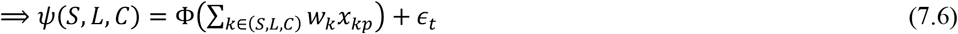

*f*_*tp*_ *i*s the observed (actual) parameter value (logarithm) at time *t, f*_*tp*_ (*t*) *i*s its time dimension decomposition, and *ψ*(*S, L, C*) *i*s its parameter dimension decomposition; *a i*s the regression intercept in the time range *T* (*t∈[0,T])*, and *ϵ*_*R*_ *i*s the regression residual at time *t*; *x*_*jt*_ *i*s the value of the *j*th time factor (*j∈[1,J])* at time *t*, and *w*_*j*_ is its regression coefficient, i.e., the weight; *x*_*kp*_ *i*s the value of the *k*th factor (*k∈(S,L,C))* in the parameter dimension, and *w*_*k*_ *i*s the regression coefficient, i.e., the weight; Θ(∙) *i*s the “overall” activation function in the time dimension, and Φ(∙) *i*s the “overall” activation function in the parameter dimension.

*f*_*tp*_ *i*s the observed (actual) parameter value (logarithm) at time *t, f*_*tp*_ (*t*) *i*s its time dimension decomposition, and *ψ*(*S, L, C*) *i*s its parameter dimension decomposition; *a i*s the regression intercept in the time range *T* (*t∈[0,T])*, and *ϵ*_*R*_ *i*s the regression residual at time *t*; *x*_*jt*_ *i*s the value of the *j*th time factor (*j∈[1,J])* at time *t*, and *w*_*j*_ *i*s its regression coefficient, i.e., the weight; *x*_*kp*_ *i*s the value of the *k*th factor (*k∈(S,L,C))* in the parameter dimension, and *w*_*j*_ *i*s the regression coefficient, i.e., the weight; Θ(·) *i*s the “overall” activation function in the time dimension and Φ(∙) *i*s the “overall” activation function in the parameter dimension; *f*_*tp*_(*t ™ τ*) *i*s the observed (actual) parameter value (logarithm) (*τ∈[1,t-1]*) value at time *t-τ*, and *w*_*k*−τ_ *i*s its autoregressive coefficient, or the weight; Γ(∙) *i*s the temporal autocorrelation “overall” activation function of *f*_*tp*_(*t*), representing a nonlinear higher-order polynomial fit of the autoregression.

The difference between Equations 6 and 7 is that LSTM includes a temporal autocorrelation of *f*_*tp*_(*t*) *i*tself, so that it is not only a sequence in the parameter dimensions but also a time series.

### 2.3 Data

We collect the following COVID-19 datasets from two sources:

1. Dataset 1: A JHU CSSE dataset (https://raw.githubusercontent.com/CSSEGISandData/COVID-19/master/csse_covid_19_data/csse_covid_19_time_series/), which tracks confirmed cases and deceased cases. We use the confirmed/deceased dataset to form training data for deep learning.
2. Dataset 2: A Tencent dataset (https://view.inews.qq.com/g2/getOnsInfo?name=disease_other), which updates daily records (confirmed, active, deceased, recovered, etc.). We use these detailed case data to construct the compartmental model.

In general, both datasets have some reporting discrepancies, with certain extreme outliers in both directions; thus, we run a 7-day moving average on the datasets to smooth out these data irregularities.

### 2.4 Modeling Methodology

We then conduct the following step-by-step operations to model the Omicron epidemic in mainland China, Hong Kong and Taiwan (steps 1 and 2) and the whole country (all steps). Fig. 1 is the flowchart to illustrate the modeling methodology.

**Fig. 1.**
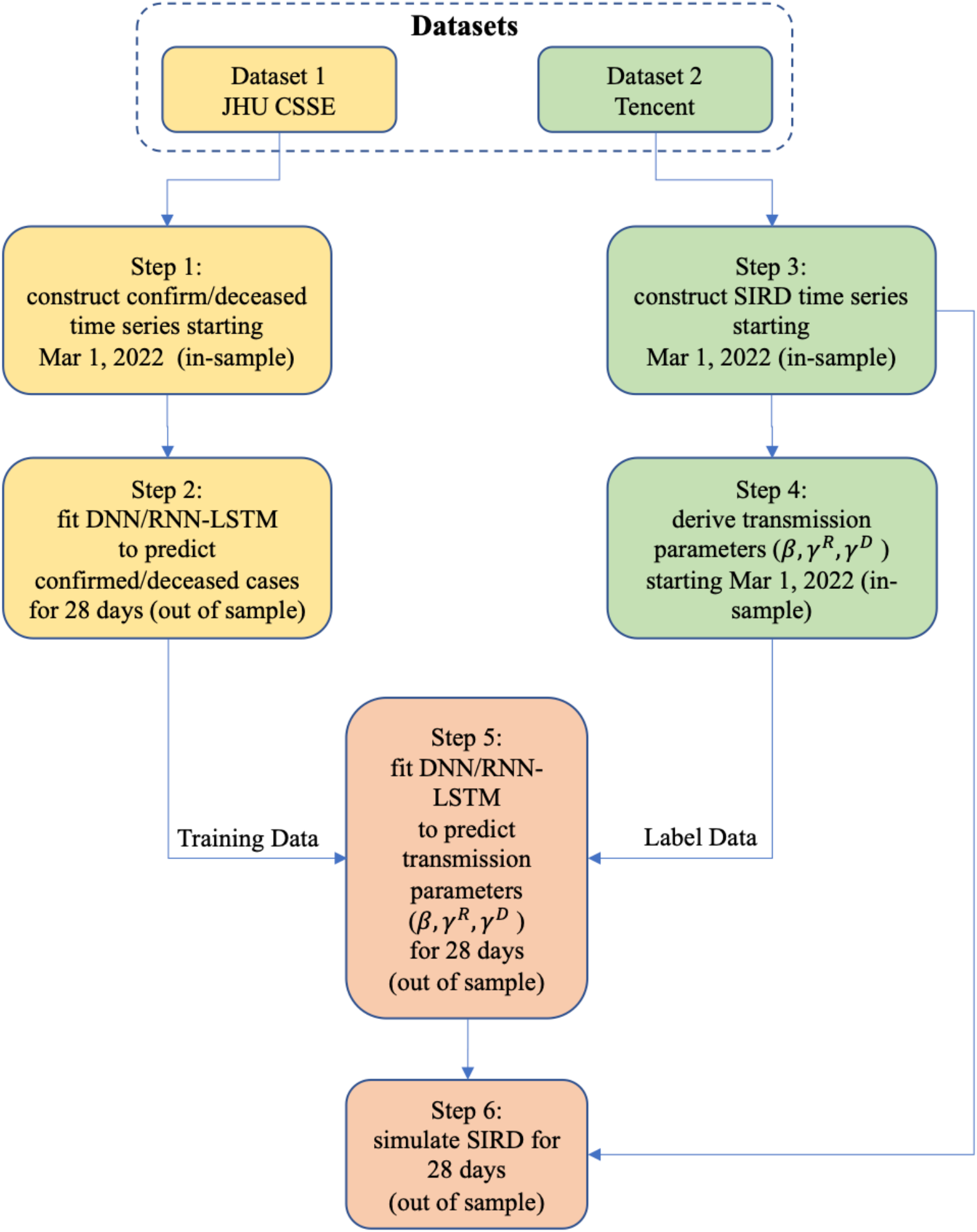
The methodology flowchart

1. We construct a confirmed/deceased time series starting from Mar 1, 2022 (in-sample) from Dataset 1. Mar 1, 2022, is generally accepted as the outbreak point of the Omicron epidemic in China [13].
2. We apply two deep learning models (DNN and LSTM) to fit the confirmed/deceased in-sample time series from Step 1 and predict the further development of confirmed/deceased cases for 28 days (out-of-sample).
3. We construct an in-sample SIRD time series starting on Mar 1, 2022, from Dataset 2.
4. We use the in-sample SIRD time series constructed in Step 3 to come up with in-sample sequences for the SIRD daily transmission parameters (*β, γ*^*p*^ *a*nd *γ*^*D*^).
5. We then use the confirmed/deceased time series (in-sample and out-of-sample) from Step 2 as training data, the in-sample *β, γ*^*p*^ *a*nd *γ*^*D*^ *s*equences from Step 4 as training labels, and apply the DNN and RNN-LSTM techniques to predict *β, γ*^*p*^ *a*nd *γ*^*D*^ *f*or 28 days (out-of-sample).
6. Finally, we use the predicted (out-of-sample) transmission parameters (*β, γ*^*p*^ *a*nd *γ*^*D*^) from Step 5 to simulate the 28-day progression (out-of-sample) of the SIRD model in a recursive manner, starting from the last timestep from the in-sample SIRD time series from Step 4.
7. We then repeat Steps 1-6 for Hong Kong and Taiwan to test the robustness of the model with data from different phases (and populations) of the epidemic.

## 3. Results

The average results of eight models (scenarios with different learning hyperparameters) based on data up to June 3, 2022 are illustrated in Figs. 2 – 4 (28-day forecast). The predicted numbers of infections, deaths and case fatality rate (CFR) are listed in Table 1.

**Table 1.** Prediction Accuracy.

|  | Predicted |  |  | Reported |  |  | Prediction Error (absolute) |  |
| --- | --- | --- | --- | --- | --- | --- | --- | --- |
|  | Infection | Death | Case Fatality Rate | Infection | Death | Case Fatality Rate | Infection | Death |
| China mainland | 768,622 | 591 | 0.0769% | 768,935 | 621 | 0.0808% | 0.04% | 5.08% |
| Hong Kong | 1,220,352 | 9,282 | 0.7606% | 1,228,002 | 9,186 | 0.7480% | 0.63% | 1.03% |
| Taiwan | 3,842,576 | 4,482 | 0.1166% | 3,669,157 | 5,595 | 0.1525% | 4.51% | 24.83% |
| China | 5,587,799 | 15,380 | 0.2752% | 5,666,094 | 15,402 | 0.2718% | 1.40% | 0.14% |

**Fig. 2.**
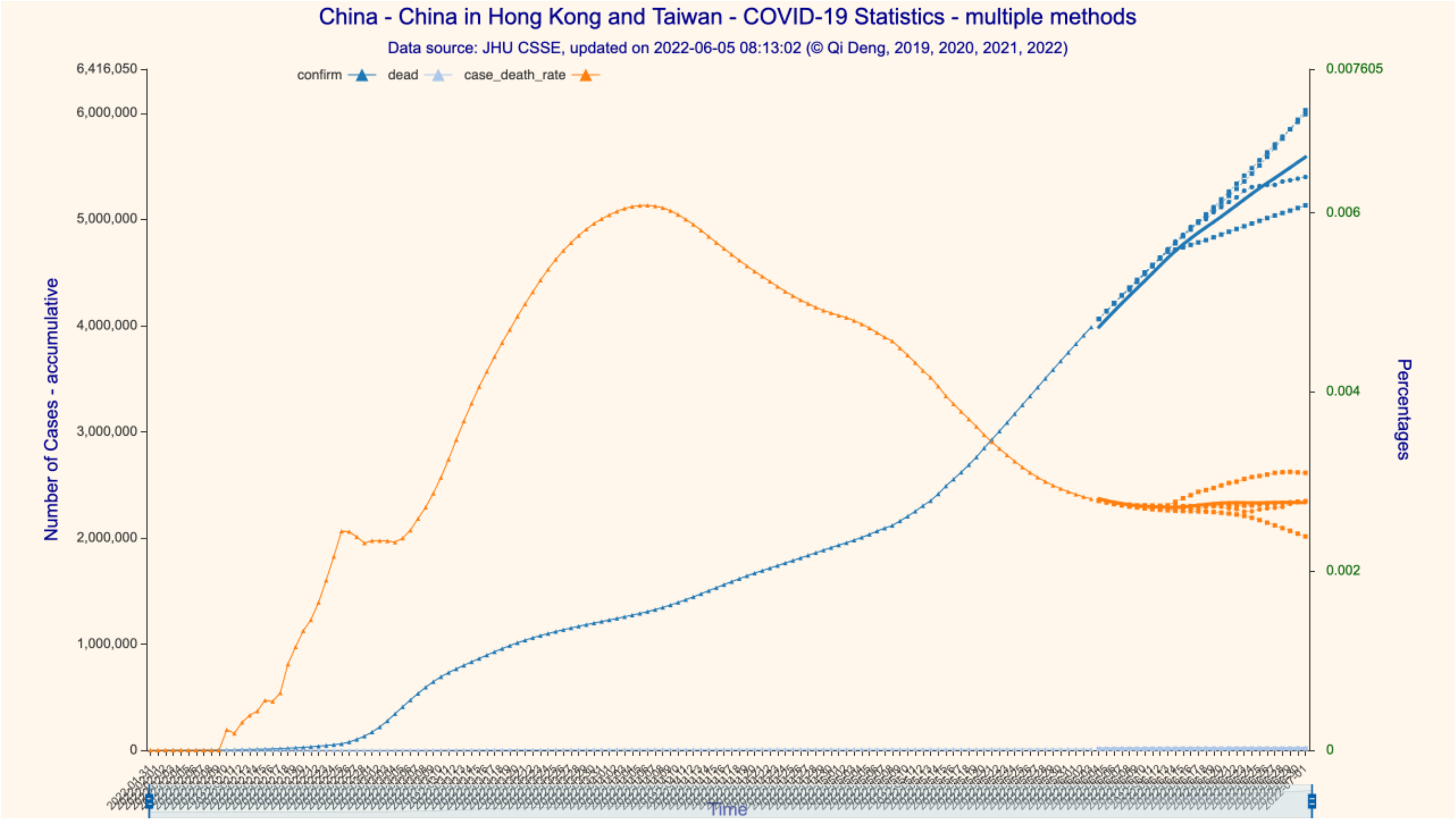
28-day forecast for the confirmed/deceased cases in China (incl. Hong Kong and Taiwan)

We predict that in the 28-day forecast period, in mainland China (excluding Hong Kong and Taiwan), the daily Omicron infection increase is between 60 and 260. On July 1, 2022, there would be 768,622 cumulative confirmed cases and 591 cumulative deceased cases, with a CFR of 0.0769%. On July 1, 2022, in Hong Kong, there would be 1,220,352 cumulative confirmed cases and 9,282 cumulative deceased cases, with a CFR of 0.7606%. We further predict that the number of daily infection increases in Hong Kong would be flat and at a very low level until at least the end of June 2022, between 350 and 1,100. On July 1, 2022, in Taiwan, there would be 3,842,576 cumulative confirmed cases and 4,482 cumulative deceased cases, with a CFR of 0.1166%. The Omicron epidemic in Taiwan is far from being over, the number of daily infection increases peaked by the end of May at approximately 83,000 and would drop to approximately 41,750 by July 1, 2022.

Since dataset 2 only provides detailed time series case data for mainland China, we are only able to construct the SIRD time series for the whole country, not for the subregions. We predict that the numbers of cumulative confirmed/deceased cases would be 5,587,799 and 15,380, respectively, with a CFR stabilizing at 0.2752% on July 1, 2022 (Figs 2 and 3). We then forecast the transmission parameters and simulate the dynamics and development of the Omicron epidemic with the SIRD time series construct (Fig 4). For the 28-day time period ending on July 1, 2022, we find that the daily infection increase has already peaked at the end of May and would drop steadily to near zero around June 14, 2022, but then rise at a low level between 150 and 4,000 afterwards.

**Fig. 3.**
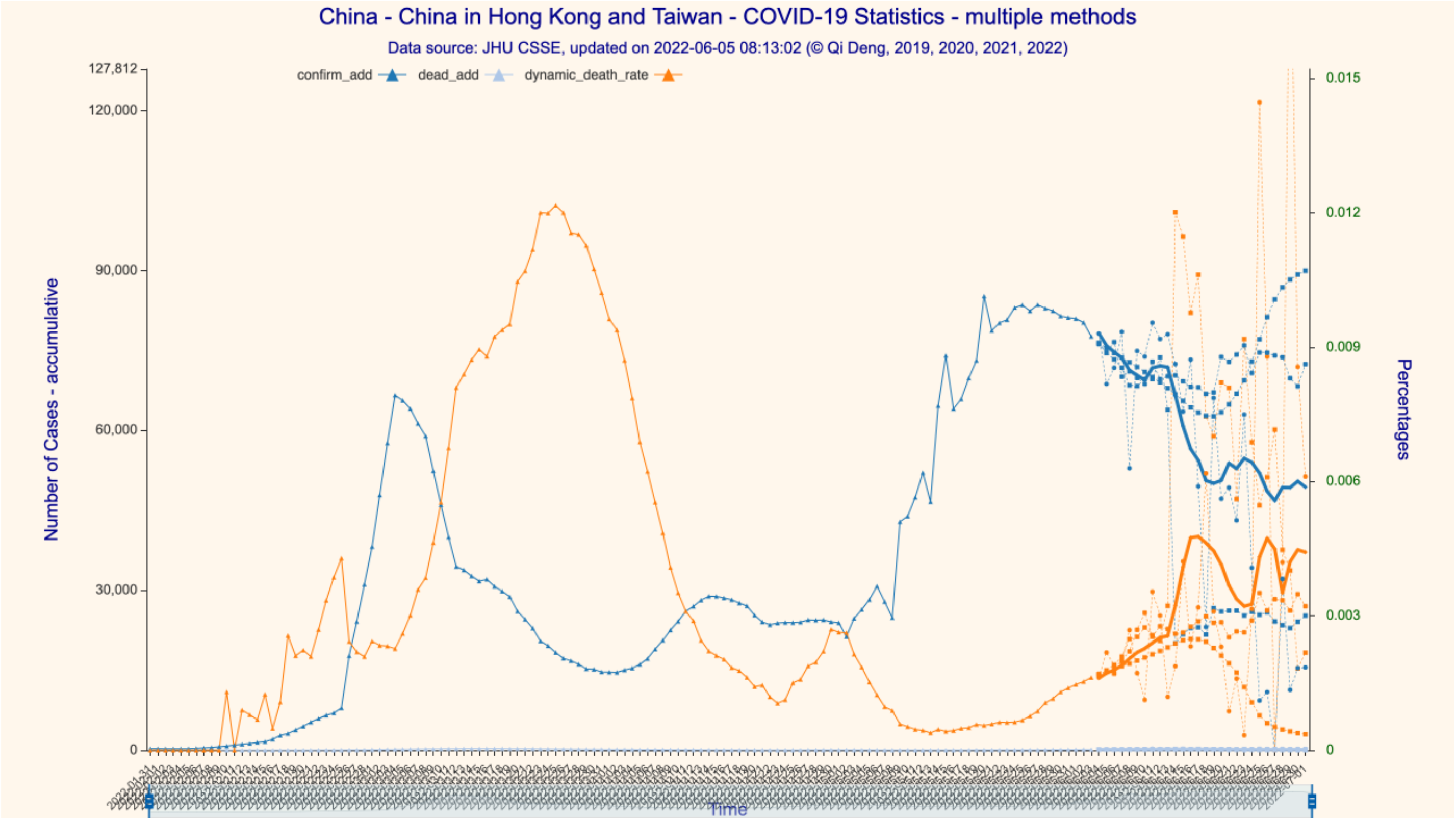
28-day forecast for the increase in confirmed/deceased cases in China (incl. Hong Kong and Taiwan)

**Fig. 4.**
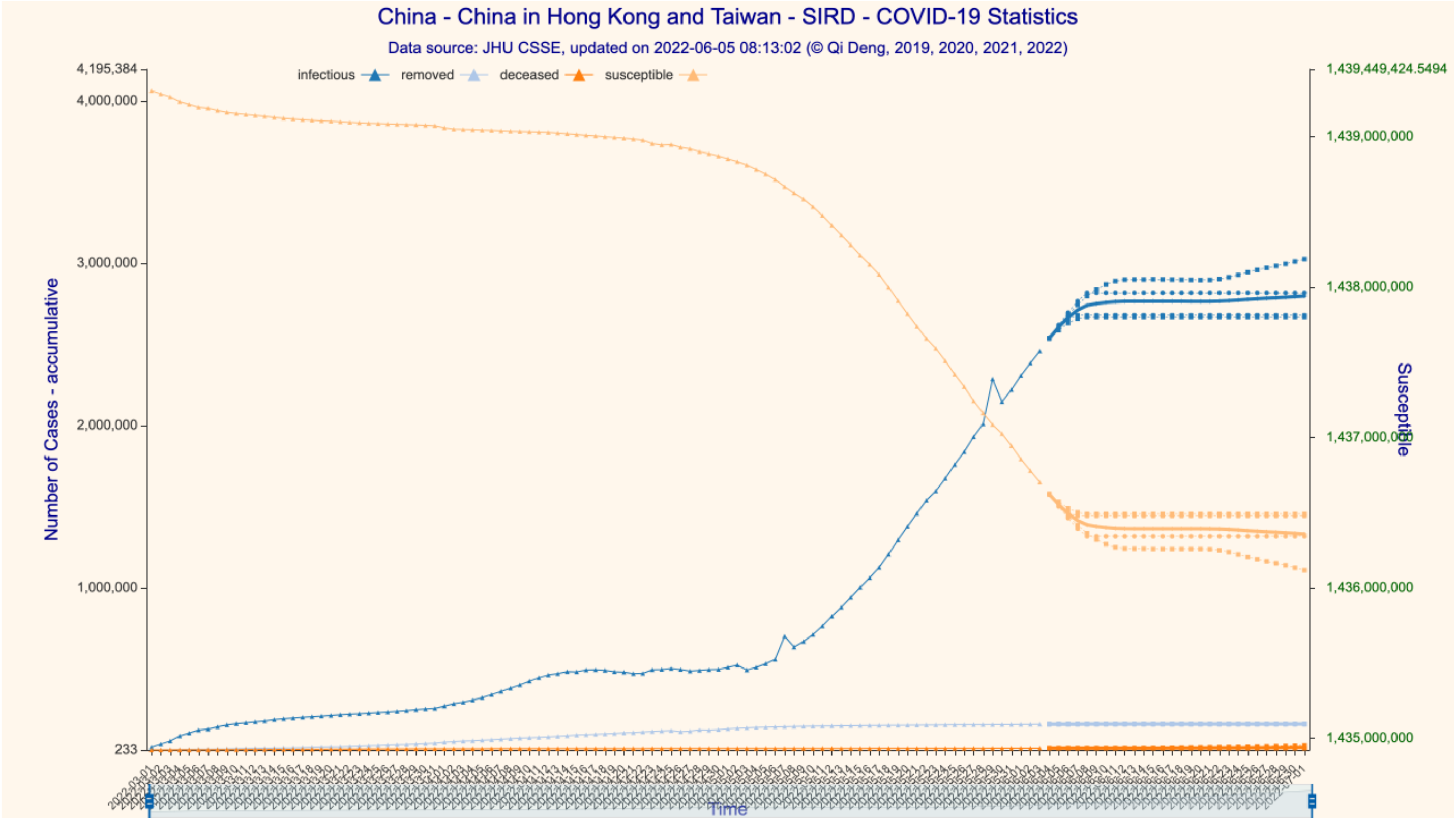
28-day SIRD simulation for cases in China (incl. Hong Kong and Taiwan)

We then compare our predictions against the reported (actually) data for the same 28-day time span (Table 1). The prediction errors of our model are at different levels of accuracy depending upon regions and case types. For the number of infections, the prediction error for mainland China is the lowest at 0.04% (99.96% accuracy), which can be explained by China’s extremely strict zero-COVID policy reducing the volatility of transmission dynamics to an extremely low level. The prediction error for Taiwan is the highest at 4.51% (95.49% accuracy), as Taiwan did not take any extra measures against COVID that would have interrupted the daily routines and movement of the populace; thus, COVID transmission essentially took its natural path. The prediction error for Hong Kong is in the middle at 0.62% (99.38% accuracy) because Hong Kong executed a somewhat middle-of-way strategy that balanced disease control and economic development. That the CFR for Hong Kong is much higher than that for Taiwan (0.7480% vs. 0.1525%) can be explained by the very low vaccination rate in Hong Kong at the time, especially among the elderly. As such, the model’s lower limit of accuracy on infection prediction is at a very respectable level of higher than 95%, and the average level of accuracy is approximately 98%.

For the numbers of deaths, the prediction errors are 5.08%, 1.03%, 24.83% and 0.14% for mainland China, Hong Kong, Taiwan and the whole country, respectively. The results suggest that the level of accuracy of death prediction is generally higher with an increased population base. That the prediction error for Taiwan is the highest (24.83%) is due to the overwhelming pressure on the island’s healthcare system. The model’s lower limit of accuracy on death prediction is higher than 75%, and the average level of accuracy is approximately 92%.

## 4. Discussion

We apply DNN and LSTM techniques to estimate the stochastic transmission parameters for a SIRD model with a discrete time series construct. We then apply DNN and LSTM deep learning techniques to fit the confirmed/deceased time series to predict further development of confirmed/deceased cases, as well as to predict the transmission parameters (*β, γ*^*p*^, *γ*^*D*^) for 28 days. Finally, we use the predicted transmission parameters to simulate the Omicron dynamics for 28 days. The average levels of prediction accuracy of the model are 98% and 92% for the numbers of infections and deaths, respectively.

With the introduction of the deep learning-enhanced compartmental model, we provide an effective and easy-to-implement alternative to prevailing stochastic parameterization. The deep learning techniques uncover hidden interconnections among observed data, which greatly reduces the dependency on data particularity. Our model demonstrates the efficacy and potential of applying deep learning methods in predicting the dynamics of infectious diseases.

## Statements and Declarations

The work was supported by Hubei University of Automotive Technology [grant number BK202209] and Hubei Provincial Bureau of Science and Technology [grant number 2023EHA018].

The authors have no relevant financial or non-financial interests to disclose.

## Data Availability

datasets extracted from JHU CSSE and Tencent

https://raw.githubusercontent.com/CSSEGISandData/COVID-19/master/csse_covid_19_data/csse_covid_19_time_series

https://view.inews.qq.com/g2/getOnsInfo?name=disease_other

## Figure Legend

[1] confirm – accumulative number of confirmed infections

[2] dead – accumulative number of death

[3] case death rate = dead/confirm, representing overall death rate

[4] confirm_add – daily increase in number of confirmed infections

[5] dead_add – daily increase in number of death

[6] dynamic_death_rate = dead_add/confirm_add, representing the trend of death rate

## Notes

### Competing Interest Statement

The authors have declared no competing interest.

### Summary of Updates

Change of author affiliation, change of abstract and content to reflect the accuracy of the model, reduce number of figures to streamline and simplify the paper.

